# Education Intervention for Evaluation and Living Donor Kidney Transplantation: A randomized trial

**DOI:** 10.64898/2026.03.10.26348081

**Authors:** Miriam Vélez-Bermúdez, Jamie M. Loor, Yuridia Leyva, L. Ebony Boulware, Yiliang Zhu, Mark L. Unruh, Emilee Croswell, Amit Tevar, Mary Amanda Dew, Larissa Myaskovsky

**Author notes:** **Corresponding author:** Miriam Vélez-Bermúdez, PhD MPH, Center for Healthcare Equity in Kidney Disease, The University of New Mexico Health Sciences Center 901 University Blvd SE STE 150, Albuquerque, NM, 87106.

## Abstract

**Key Points:**

- In a randomized trial, an educational booklet and video did not increase evaluation completion or living donor kidney transplant receipt.
- For patients who received the booklet and video intervention, experiencing discrimination in healthcare reduced evaluation completion.
- Long-term follow-up and a large sample size yielded sufficient power to validate a true null effect of the intervention on key outcomes.

**Background:** Kidney transplantation (KT) evaluation is a complex, lengthy process; and living donor KT (LDKT) is the optimal treatment for kidney failure. Interventions at the start of evaluation may improve evaluation completion and LDKT rates. This study tested whether (a) an educational booklet and video (the “Talking About Living Kidney donation” [TALK] intervention) increased evaluation completion and LDKT when delivered under a streamlined KT evaluation program; and (b) if no effects found, explore differential effects by psychosocial/sociocultural factors (e.g., healthcare-related discrimination).

**Methods:** We conducted a randomized-controlled trial of the TALK intervention using permuted block randomization at an urban transplant center. Participants were enrolled 05/2015-06/2018; follow-up through 08/2022. Staff were blinded to block size, not allocation. Fine-Gray proportional hazards models examined intent-to-treat and per-protocol approaches. Primary outcomes were the cumulative incidence of evaluation completion and LDKT receipt. We explored interaction analyses by psychosocial/sociocultural factors and TALK-assignment.

**Results:** Among 1108 participants (574 [52%] TALK, 534 [48%] No-TALK; median age: 59.13 [IQR: 48.92-67.10]; 243 [22%] Black, 783 [71%] White, 82 [7%] Other; 695 [63%] male), TALK did not significantly improve evaluation completion (sub-distribution hazard [SHR]=1.06; 95% CI: 0.92-1.22) or LDKT receipt (SHR=0.83; 95% CI: 0.55-1.25) in intent-to-treat and per-protocol analyses. In exploratory per-protocol analyses, discrimination significantly modified the effect of TALK on evaluation completion (SHR=0.42; 95% CI: 0.29-0.61). The “No-Discrimination” TALK participants had greater evaluation completion than No-TALK (SHR=1.32; 95% CI: 1.10-1.58), but the “Discrimination” TALK participants had lower evaluation completion than No-TALK (SHR=0.56; 95% CI: 0.41-0.77).

**Conclusions:** Despite streamlined care, TALK did not improve evaluation completion or LDKT rates. A significant interaction in the per-protocol analyses for evaluation completion suggests prior healthcare-related discrimination may limit educational intervention effectiveness. Future studies should explore approaches that address systemic barriers and complement, rather than rely on, educational strategies to promote LDKT (ClinicalTrials.gov Identifier: NCT02342119).

## Introduction

Kidney transplantation (KT) reduces mortality, improves quality of life, and costs less than dialysis for patients with kidney failure.^1–4^ These advantages are greater with living donor KT (LDKT) versus deceased donor KT (DDKT).^1,4–6^ However, kidney failure patients face multiple barriers to LDKT. Younger age, lower income, being underinsured or uninsured, less KT knowledge, and more LDKT-related concerns are associated with lower probability of LDKT than DDKT.^7–11^ Among LDKT recipients, non-Hispanic Whites comprise the majority, while non-Hispanic Blacks, Latine/Hispanics, Native Americans and Alaskan Natives are underrepresented.^12–14^

Before patients with kidney failure may be considered for any KT, they must complete an often burdensome, multi-step evaluation process involving multiple physicians and tests.^15^ This evaluation process represents a critical period for interventions to improve the likelihood of KT waitlisting and LDKT access, as one cannot be waitlisted or transplanted without being evaluated.^16–18^

Thus, we conducted a randomized-controlled trial (RCT) initiated at evaluation start, within a high-quality care context that streamlined evaluation for all patients, dubbed Kidney Transplant Fast Track (KTFT).^19,20^ This RCT focused on a novel deployment of the Talking About Living Kidney donation (TALK) educational materials. TALK, a culturally-tailored, psychoeducational booklet and video developed by Boulware and colleagues,^21,22^ was informed by racially- and ethnically-diverse patients and families who identified barriers to LDKT, and aimed to enhance patient and family consideration of LDKT by addressing common misconceptions about the KT process and enhancing trust.^21^ We predicted that testing TALK within the KTFT program would create ideal clinical conditions in which the intervention could have the greatest potential impact in promoting evaluation completion and LDKT receipt.

Prior TALK intervention studies focused on KT waitlisted patients, and paired the intervention with resource-intensive strategies like social worker involvement and financial assistance to potential living donors.^22–24^ These trials improved some intermediate outcomes, like for example, increased discussions with family and physician(s) about LDKT,^22^ but the trials did not demonstrate increased LDKT rates.^24^ Because TALK was designed to support patient-family consideration of LDKT,^22^ we introduced TALK at *evaluation initiation*—prior to waitlisting, while there are still opportunities for decision-making and discussions with potential living donors, and when intervening could potentially improve both evaluation completion *and* LDKT access. Additionally, assessing both outcomes allowed us to capture TALK’s impact at the beginning of the patient journey, and at an optimal endpoint (i.e., LDKT receipt).

Another innovation of our trial is testing TALK as an education-only intervention. Previous studies required resource-intensive supportive strategies (e.g., social worker involvement),^24^ which limit feasibility and scalability in resource-constrained settings. Testing TALK effectiveness without layering resource-intensive components could demonstrate its feasibility for broader implementation.

Our trial further extended prior work by considering psychosocial and sociocultural factors that may modify TALK effectiveness. Previous TALK studies considered some covariates (e.g., race/ethnicity, family function),^22,24^ but did not examine if intervention effects varied by any psychosocial or sociocultural factors. Factors including religiosity or sociocultural variables like patients’ experiences of healthcare-related discrimination, racism, medical mistrust, or trust in physicians are well-documented as predictors of health behaviors and outcomes;^25–33^ and our prior work found that these factors changed after participants underwent KTFT’s concierge-based approach to evaluation.^34^ Given their relevance in KT-related contexts, as well as mixed findings in prior TALK trials,^22–24^ we planned to examine how these factors influenced intervention effectiveness due to their potential to shape patients’ ability to complete evaluation or pursue LDKT.

Thus, our RCT tested the effectiveness of providing TALK education materials at the start of evaluation, among patients receiving streamlined, high-quality care, which we expected would reduce barriers that may otherwise limit the intervention’s effect. We tested two hypotheses: 1) given TALK’s potential to enhance patient engagement and proactivity,^24^ TALK would increase patients’ evaluation completion rates; and, 2) contingent on evaluation completion and subsequent waitlisting, TALK would increase LDKT receipt. As an exploratory aim, if no effects were found for these hypotheses, we planned to examine whether intervention effects on both outcomes differed by psychosocial or sociocultural factors, given mixed findings from prior TALK studies,^22–24^ and past evidence of these factors’ ability to predict KT access and related outcomes.^25–33^ These exploratory analyses were intended to generate hypotheses on differential effects of TALK.

## Methods

### Study Design and Sample

This RCT (ClinicalTrials.gov identifier: NCT02342119, “Increasing Equity in Transplant Evaluation and Living Donor Kidney Transplantation”) was conducted at the University of Pittsburgh Medical Center Starzl Transplant Institute. We recruited from a prospective cohort of patients undergoing KT evaluation from 05/2015-06/2018. Patient evaluation completion and receipt of LDKT were followed via medical records through 08/2022. All participants underwent KTFT (see Bornemann et al. 2017 for detailed protocol).^19^ The study follows Consolidated Standards Of Reporting Trials reporting guidelines (see Supplemental Table 1).^35^

Study eligibility requirements included patients age 18 years or older, English-speaking, with no cognitive/sensory impairments that would prevent participation, no prior KT, and not on another center’s KT waitlist.^19^ Participants provided verbal consent and completed baseline telephone interviews before their first KT evaluation clinic appointment (Figure 1 shows patient flow). This study was approved by the Institutional Review Boards at the University of Pittsburgh (PRO09060113) and the University of New Mexico (17–084), and both institutions signed a data use agreement. Study conduct was in accordance with the Declaration of Helsinki and consistent with the Principles of the Declaration of Istanbul as outlined in the ‘Declaration of Istanbul on Organ Trafficking and Transplant Tourism.’

**Figure 1.**
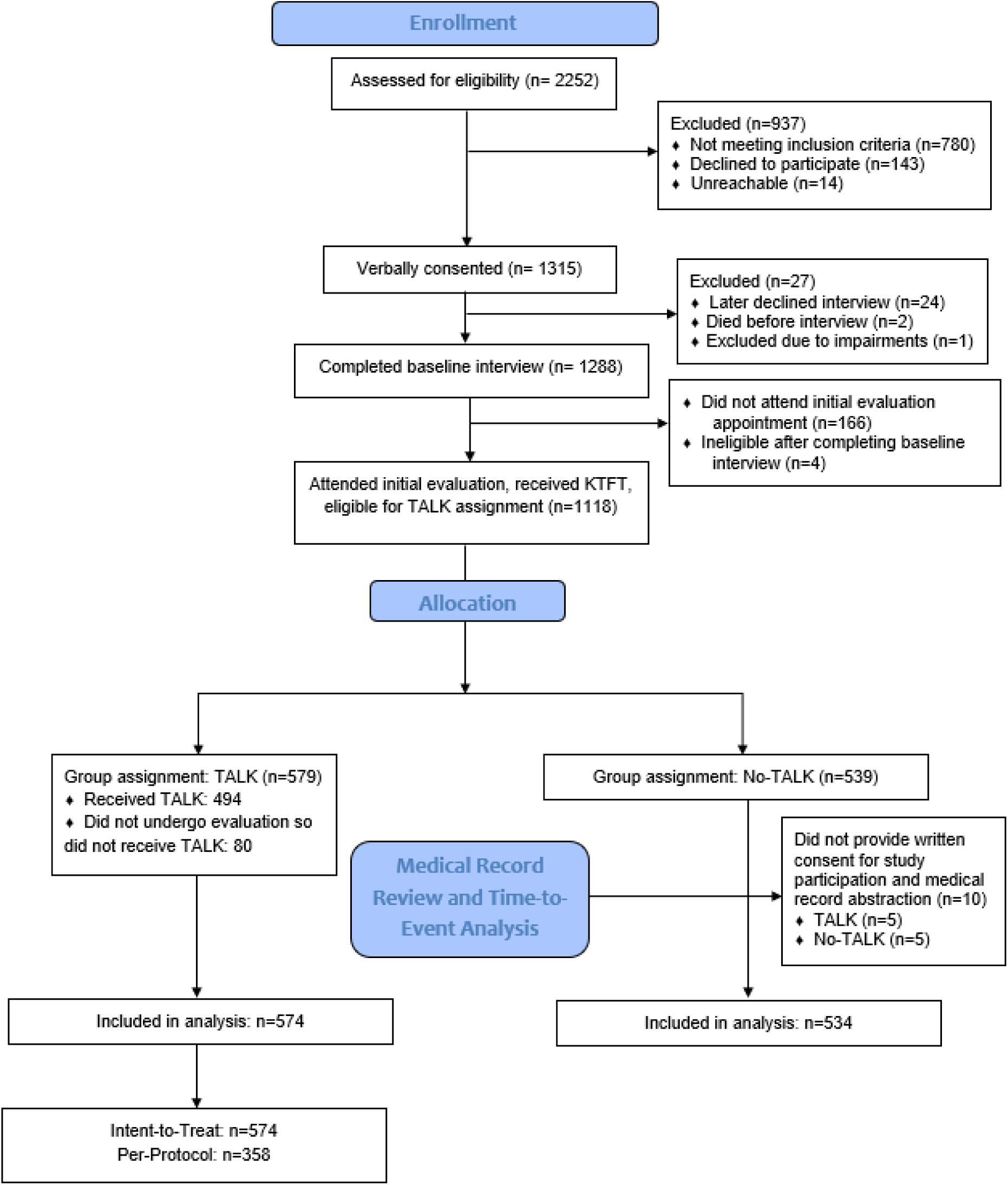
Flowchart: Cohort enrolled in intervention

### Randomization

Participants were randomized to the TALK intervention (“TALK”) or usual care (“No-TALK”). We used permuted block-randomization with an initial allocation ratio of 1:1. The schedule, with block sizes of 2-10 generated by a biostatistician at study inception, was kept locked in the principal investigator’s office cabinet. Research assistants enrolling participants were blinded to block *size* but not *allocation*. During recruitment, 80 participants randomized to TALK did not proceed with evaluation, thus never received TALK materials. To ensure sufficient intervention exposure among TALK participants, the allocation ratio was adjusted in 08/2017 to 1.22:1 (i.e., 0.55 probability of receiving TALK).

### TALK Intervention

Participants randomized to TALK were provided with the TALK booklet and video (©2008, Johns Hopkins University and National Kidney Foundation of Maryland).^19,21^ The booklet was written at a 4^th^-6^th^ grade reading level. It covered LDKT eligibility, evaluation procedures/concerns, donor selection process, and donor/recipient surgical procedures.^21^ The video was available on DVD or online, featuring diverse patients and families sharing their experiences with LDKT and its impact. The video also featured physicians and social workers, and addressed patients’ mistrust in the KT process.^21^

After two weeks, study coordinators called TALK participants to encourage review of materials and answer any questions about the material content. From this call, study coordinators entered into the study database whether patients had reviewed either or both the TALK booklet and video.^19^

### Power

Power was estimated at the study design stage to guide recruitment goals using a prior study of 1152 KT candidates within the same clinic.^7,20^ In that prior cohort, approximately 57% (n=652) reached active waitlisting, and about one-third (n=396) ultimately received a KT, including 117 confirmed LDKTs.^7,20^ Because LDKT receipt is the least common outcome along the KT trajectory,^14^ we powered the study based on this event, ensuring an adequately sized sample. Assuming a conservative baseline proportion of LDKT rates between TALK-assignment, a total sample size of 876 (1:1 allocation, 438 per arm) was estimated to generate 80% power (α=.05) to detect a 9% absolute difference in LDKT rates between groups. Although this power calculation was based on a binary LDKT receipt outcome, the primary analyses used competing risk time-to-event models, which leveraged follow-up time and censoring to enhance observed power.

### Study Measures

**Independent Variables:** The primary independent variable was randomization to TALK (“TALK” versus “No-TALK”). Other *pre-specified variables* forced into adjusted models included:

- **Healthcare-related sociocultural factors**: experience of discrimination in healthcare,^26,36^ perceived racism in healthcare,^37^ medical mistrust in healthcare systems,^37–39^ and trust in physician(s)^40^
- **Psychosocial and cultural factors** previously associated with KT-related outcomes, including family loyalty, health literacy,^41^ religiosity, and religious objection to LDKT^42^
- **Race and ethnicity**, because TALK was designed with racially- and ethnically-diverse patients in mind (Supplemental Table 2).^21^

*Candidate Covariates*: Baseline covariates considered for model inclusion were **demographic characteristics** (e.g., age); **medical factors** (e.g., dialysis type); **donation preference and recruitment** (e.g., DDKT/LDKT preference); **transplant knowledge and concerns,** and additional **psychosocial factors** (e.g., social support; Supplemental Table 2).

**Outcome Variables** were the cumulative incidence of 1) evaluation completion, and 2) LDKT receipt.

### Statistical Analysis

We examined data for missingness, distributional characteristics, and analytic assumptions (e.g., normality, skewness, outliers), and made appropriate adjustments (e.g., transformations/categorization). For example, we dichotomized the healthcare discrimination measure into “Ever” and “Never” based on its skewness.^36^ Frequencies of missing cases for each variable are reported in Table 1, footnote a. We calculated frequencies and percentages for categorical variables, and medians and interquartile ranges for continuous variables.

**Table 1.** Characteristics of participants at baseline: Total sample, and by Intervention Group (TALK or No-TALK).

| Characteristic <sup>a</sup> | Total (N=1108) | TALK (N=574) | No-TALK (N=534) | Test Statistic $\chi^2$ (df) or z statistic | p-value <sup>c, d</sup> |
| --- | --- | --- | --- | --- | --- |
| **Demographics factors** |  |  |  |  |  |
| Race and ethnicity: n (%) |  |  |  | 0.02 (2) | .992 |
| Non-Hispanic White | 783 (71) | 405 (71) | 378 (71) |  |  |
| Non-Hispanic Black | 243 (22) | 126 (22) | 117 (22) |  |  |
| Other race or ethnicity <sup>b</sup> | 82 (7) | 43 (7) | 39 (7) |  |  |
| Age at T1 completion: Median (IQR) | 59.13 (48.92, 67.10) | 59.09 (47.80, 66.96) | 59.20 (49.84, 67.45) | -0.86 | .389 |
| Gender: n (%) |  |  |  | 1.00 (1) | .317 |
| Men | 695 (63) | 352 (61) | 343 (64) |  |  |
| Women | 413 (37) | 222 (39) | 191 (36) |  |  |
| Educational level (≤ High School): n (%) | 513 (46) | 270 (47) | 243 (46) | .26 (1) | .609 |
| Family Income (< \$50,000): n (%) | 772 (72) | 408 (74) | 364 (71) | .87 (1) | .351 |
| Marital Status (married/domestic partnership): n (%) | 531 (48) | 261 (45) | 270 (51) | 2.87 (1) | .090 |
| Employment status (part- or full-time employed): n (%) | 268 (24) | 131 (23) | 137 (26) | 1.21 (1) | .271 |
| Number in social network: Median (IQR) | 19 (11, 32.50) | 19 (11, 34) | 19 (11, 31) | .23 | .819 |
| Log (Number in social network): Median (IQR) | 2.94 (2.40, 3.48) | 2.94 (2.40, 3.53) | 2.94 (2.40, 3.43) | .23 | .819 |
| Insurance: n (%) |  |  |  | **8.03 (2)** | **.018** |
| Private only | 202 (18) | 88 (15) | 114 (21) |  |  |
| Public only | 478 (44) | 265 (46) | 213 (40) |  |  |
| Both Private and Public | 428 (39) | 221 (39) | 207 (39) |  |  |
| **Medical factors** |  |  |  |  |  |
| Type of Dialysis: n (%) |  |  |  | 1.11 (2) | .573 |
| Hemodialysis | 568 (51) | 296 (52) | 272 (51) |  |  |
| Peritoneal dialysis | 128 (12) | 71 (12) | 57 (11) |  |  |
| No dialysis | 412 (37) | 207 (36) | 205 (38) |  |  |
| Dialysis duration: n (%) |  |  |  | 0.62 (3) | .891 |
| No time on dialysis | 392 (35) | 201 (35) | 191 (36) |  |  |
| <1 year on dialysis | 419 (38) | 220 (38) | 199 (37) |  |  |
| 1-5 years on dialysis | 236 (21) | 124 (22) | 112 (21) |  |  |
| >5 years on dialysis | 61 (6) | 29 (5) | 32 (6) |  |  |
| Body Mass Index (BMI): Median (IQR) | 29.57 (25.57, 34.97) | 29.48 (25.54, 34.31) | 29.69 (25.64, 35.54) | -0.95 | .345 |
| Burden of kidney disease: Median (IQR) | 4 (3, 4.67) | 4 (3, 4.67) | 4 (3, 4.67) | 0.79 | .429 |
| Charlson Comorbidity Index (CCI): Median (IQR) | 4 (3, 5) | 4 (3, 5) | 4 (3, 5) | 1.47 | .142 |
| **Psychosocial Characteristics** |  |  |  |  |  |
| Social support: Median (IQR) | 43 (37, 47) | 43 (38, 47) | 43 (37, 46) | 0.96 | .335 |
| Anxiety (≥ moderate): n (%) | 54 (5) | 23 (4) | 31 (6) | 1.93 (1) | .165 |
| Depression (≥ moderate): n (%) | 71 (6) | 30 (5) | 41 (8) | 2.77 (1) | .096 |
| **Transplant Knowledge and Concerns** |  |  |  |  |  |
| Transplant knowledge: Median (IQR) | 11 (9, 13) | 11 (9, 13) | 11 (9, 13) | 0.10 | .920 |
| Number of learning activities: Median (IQR) | 2 (1, 3) | 2 (1, 3) | 2 (1, 3) | 0.06 | .949 |

| Characteristic <sup>a</sup> | Total (N=1108) | TALK (N=574) | No-TALK (N=534) | Test Statistic<br>x<sup>2</sup> (df) or z statistic | p-value <sup>c, d</sup> |
| --- | --- | --- | --- | --- | --- |
| Total hours of learning activities: Median (IQR) | 4 (2, 9) | 4 (2, 10) | 4 (2, 8) | 1.46 | .144 |
| Transplant concerns: Median (IQR) | 87 (78, 96) | 88 (78, 96) | 87 (78, 96) | -0.12 | .906 |
| **Donor Recruitment and Preference variables** |  |  |  |  |  |
| Have a living donor at baseline (Yes): n (%) | 484 (44) | 255 (44) | 229 (43) | 0.21 (1) | .644 |
| Donor Preference: n (%) |  |  |  | 1.59 (2) | .451 |
| Deceased donor | 326 (29) | 172 (30) | 154 (29) |  |  |
| Living donor | 746 (67) | 380 (66) | 366 (69) |  |  |
| Unable to answer | 36 (3) | 22 (4) | 14 (3) |  |  |
| Willing to accept LDKT volunteer (yes): n (%) | 969 (89) | 501 (88) | 468 (89) | .17 (1) | .683 |
| Willing to ask for LDKT volunteer (yes): n (%) | 554 (52) | 282 (51) | 272 (53) | .25 (1) | .615 |
| **Survey measures included in all adjusted modeling** |  |  |  |  |  |
| Any experience of discrimination (Yes): n (%) | 335 (30) | 179 (31) | 156 (29) | 0.51 (1) | .475 |
| Perceived racism: Median (IQR) | 2.25 (2, 2.75) | 2.25 (2, 3) | 2.25 (2, 2.75) | 1.25 | .211 |
| Medical mistrust: Median (IQR) | 3 (2.57, 3.57) | 3.14 (2.57, 3.57) | 3 (2.57, 3.43) | **1.98** | **.048** |
| Trust in physician: Median (IQR) | 4 (3.64, 4.45) | 3.91 (3.55, 4.45) | 4 (3.73, 4.45) | **-2.04** | **.041** |
| Family loyalty: Median (IQR) | 3.19 (2.81, 3.56) | 3.19 (2.81, 3.60) | 3.25 (2.81, 3.56) | 0.06 | .952 |
| Religious objection to LDKT (Yes): n (%) | 798 (73) | 412 (73) | 386 (73) | 0.07 (1) | .789 |
| Religiosity: Median (IQR) | 7 (4.5, 9) | 7 (5, 9) | 7 (4.5, 9) | 1.13 | .256 |
| Health Literacy: Median (IQR) | 4 (3, 4.67) | 4 (3, 4.67) | 4 (3.33, 4.67) | -0.47 | .642 |

TALK: Talking About Living Kidney Donation
<sup>a</sup> Missing: n=40 for family income (n=19 TALK, n=21 no-TALK); n=1 for burden of kidney disease (n=1 TALK); n=2 having a living donor at baseline (n=2 no-TALK); n=36 donor recruitment and preference (n=22 TALK, n=14 no-TALK); n=16 for willing to accept a living donor (n=7 TALK, n=9 no-TALK); n=43 for willing to ask for a living donor (n=24 TALK, n=19 no-TALK); n=8 for perceived racism in healthcare (n=4 TALK, n=4 no-TALK); n=4 for medical mistrust (n=3 TALK, n=1 no-TALK); n=1 family loyalty (n=1 TALK); n=15 for religious objection to LDKT (n=7 TALK, n=8 no-TALK); n=4 religiosity (n=1 TALK, n=3 no-TALK).
<sup>b</sup> Due to the small number of participants who did not indicate “Black” or “White” race, we collapsed all other race and ethnicities into the category, “Other race or ethnicity.” Participants in the “Other race or ethnicity” group were American Indian/Native American (n=5, 0.45%), Asian (n=7, 0.63%), Hispanic/Latine (n=24, 2.17%), Native Hawaiian/Pacific Islander (n=1, 0.09%), and Mixed race or Other (n=45, 4.06%).
<sup>c</sup> Pearson's Chi-square Test for categorical variables; Wilcoxon Rank Sum Test for continuous variables.
<sup>d</sup> Bolded test statistic and p-values indicate statistical significance between TALK-assignment ( $p \leq .05$ ).

Differences in patient characteristics by TALK-assignment were calculated using Pearson’s Chi-square Tests or Wilcoxon Rank Sum Tests for categorical and continuous variables, respectively (Table 1).

#### Fine-Gray Proportional Hazards Models

We performed Fine-Gray competing risk modeling with cumulative incidence functions to determine TALK effectiveness on increasing evaluation completion and LDKT receipt, first examining unadjusted models, then models with adjustments. We chose Fine-Gray competing risk survival models rather than Cox models without competing risk, because the latter would consider terminal events to be non-informatively censored.^43^ Unlike non-terminal events (e.g., temporarily pausing evaluation), terminal events like death are distinct outcomes that preclude the possibility of evaluation completion or LDKT receipt.

To determine the effect of TALK on likelihood of evaluation completion during follow-up, we defined time starting from patients’ evaluation initiation to its completion. If evaluation was closed before completion, patients were censored at the closure date. If patients later restarted evaluation, they remained censored in analysis to capture TALK’s effectiveness on patients’ first evaluation attempt. We treated death as a competing risk event.

To determine the effect of TALK on likelihood of LDKT during follow-up, we defined time starting from active waitlisting to LDKT receipt. We treated death or DDKT as competing risk events. Patients who remained on the waitlist at the end of the study, or were removed from the waitlist for reasons other than the primary or competing risk events, were censored.

Additionally, we explored whether TALK-assignment differed in either outcome across levels of the nine forced-in demographic, sociocultural, and psychosocial variables (i.e., interactions) in separate models, all of which were adjusted with the same set of covariates as the main effect models.

#### Multiple Comparison Correction

For the exploratory interaction analyses, we addressed the risk of Type I error from multiple testing by applying the Benjamini-Hochberg false discovery rate correction.^44^ This correction was applied to the omnibus p-values corresponding to the interaction terms between TALK-assignment (TALK v. No-TALK) and each of the nine forced-in variables.

#### Intent-to-Treat and Per-Protocol Approaches

Under the intent-to-treat approach, all randomized participants who provided written consent were included in our analysis,^45^ regardless of whether they completed evaluation, or received the TALK materials (if assigned to TALK). Specifically, the 80 participants assigned to TALK who never received the education materials were included in this analysis. In addition to the intent-to-treat approach, we conducted a per-protocol analysis, retaining participants in the TALK group who self-reported reviewing the TALK booklet or video at their two-week follow-up call; we considered self-reports of reviewing the TALK booklet or video as *adherence* to the intervention.

Conducting both intent-to-treat and per-protocol analyses allowed us to assess TALK’s effectiveness under real-world conditions while also evaluating outcomes among participants who engaged with TALK as intended.^45^

#### Model Selection and Assumptions

For covariate-adjusted modeling, we applied the least absolute shrinkage and selection operator (LASSO) procedure to the candidate covariates, while holding constant the forced-in prespecified key variables (i.e., TALK assignment, experiences of discrimination in healthcare, perceived racism in healthcare, medical mistrust, trust in physicians, family loyalty, health literacy, religious objection to LDKT, religiosity, and race/ethnicity). Although randomization addresses confounding, covariate adjustment was employed to account for factors that may independently influence outcomes. Rather than including all candidate covariates in a single fully-adjusted model, we used LASSO to manage the large variable set (see Table 1). By applying regularization and shrinkage techniques, LASSO retains a parsimonious subset of predictors, effectively reducing the risk of overfitting that could stem from including less contributing covariates.^46,47^ The LASSO procedure was conducted for each outcome and accounted for competing risks through the “fastcmprsk” package in R.^48^ For each final model (comprised of the forced-in and LASSO-selected variables), we conducted the ‘proportionality of hazards’ test to confirm the assumption was met.^49^

## Results

### Baseline Characteristics

Table 1 shows baseline characteristics for the total sample (N=1108), TALK (n[%]=574[52]), and No-TALK (n[%]=534[48]) groups. Participants were predominantly non-Hispanic White (n[%]=783[71]), male (n[%]=695[63]), with a median age of 59.13 (IQR: 48.92-67.10). TALK participants were more likely to have public insurance (n[%]=265[46]) than No-TALK participants (n[%]=213[40]; χ^2^(2)=8.03, *p*=.018), but did not statistically differ on any other demographic or medical characteristic. Variables had low levels of missingness, with no systematic missingness pattern by group assignment.

In this sample, 810 participants (73%) completed evaluation (TALK=415 [72%]; No-TALK=395 [74%]) with a median follow-up of 69.5 days from evaluation start (IQR: 25, 171). A total of 108 participants (23%) received LDKT (TALK=51 [21%]; No-TALK=57 [24%]) with a median follow-up of 312 days from active waitlisting (IQR: 117, 575). Detailed counts of primary events, censored observations, competing events, and missing data for both intent-to-treat and per-protocol samples are provided in Supplemental Tables 3-4.

In the two-week follow-up, TALK participants (n=574) reported the following: 228 (40%) reviewed the booklet *and* video, 106 (18%) reviewed only the booklet, and 24 (4%) reviewed only the video. This combined total (n=358, 62%) comprised the TALK group in the per-protocol analysis. Of the remainder, n=136 received TALK but did not review the materials, and n=80 did not receive the materials.

### Fine-Gray Proportional Hazard Models

Unadjusted models indicate no effect of TALK versus No-TALK on evaluation completion (sub-distribution hazard ratio [SHR]=1.01; 95% CI: 0.88-1.16; *p*=.875; Table 2) or LDKT receipt (SHR=0.82; 95% CI: 0.56-1.20; *p*=.308; Table 2). Results from intent-to-treat and per-protocol approaches are presented side-by-side in Tables 2-3. We describe intent-to-treat findings unless explicitly noted.

**Table 2.** Fine-Gray proportional sub-distribution hazards models, examining main effects in unadjusted and adjusted models on outcomes of evaluation completion (competing event: death) and LDKT receipt (competing events: DDKT, death). Intent-to-Treat and Per-Protocol approaches are shown below.

| Variables | Evaluation Completion<sup>a</sup> |  |  |  | LDKT Receipt<sup>b</sup> |  |  |  |
| --- | --- | --- | --- | --- | --- | --- | --- | --- |
|  | Intent-to-Treat |  | Per-Protocol |  | Intent-to-Treat |  | Per-Protocol |  |
|  | SHR<br>(95% CI) | p-value | SHR<br>(95% CI) | p-value | SHR<br>(95% CI) | p-value | SHR<br>(95% CI) | p-value |
| **Unadjusted Model** |  |  |  |  |  |  |  |  |
| Group |  |  |  |  |  |  |  |  |
| TALK Assignment (TALK group)<br>(Reference: No-TALK) | 1.01<br>(0.88-1.16) | .875 | 1.00<br>(0.86-1.17) | .973 | 0.82<br>(0.56-1.20) | .308 | 0.74<br>(0.49-1.11) | .144 |
| **Adjusted Model** |  |  |  |  |  |  |  |  |
| Group |  |  |  |  |  |  |  |  |
| TALK Assignment (TALK group)<br>(Reference: No-TALK) | 1.06<br>(0.92-1.22) | .408 | 1.05<br>(0.90-1.23) | .539 | 0.83<br>(0.55-1.25) | .380 | 0.71<br>(0.45-1.11) | .134 |
| Demographic, Sociocultural, and Psychosocial Factors included in all adjusted modeling |  |  |  |  |  |  |  |  |
| Race and ethnicity<br>(Reference: Non-Hispanic White) |  |  |  |  |  |  |  |  |
| Non-Hispanic Black | 0.87<br>(0.70-1.07) | .192 | 0.88<br>(0.70-1.11) | .284 | 0.61<br>(0.28-1.35) | .222 | 0.62<br>(0.27-1.44) | .268 |
| Other race or ethnicity | 0.84<br>(0.64-1.12) | .240 | 0.92<br>(0.67-1.28) | .645 | **0.35**<br>**(0.15-0.84)** | **.019** | **0.37**<br>**(0.14-0.98)** | **.046** |
| Experiences of discrimination in healthcare<br>(Reference: Never experienced) | 1.17<br>(0.98-1.40) | .078 | 1.11<br>(0.90-1.36) | .322 | 0.90<br>(0.52-1.58) | .721 | 0.96<br>(0.53-1.73) | .883 |
| Perceived racism in healthcare | 0.97<br>(0.88-1.07) | .567 | 0.92<br>(0.82-1.03) | .153 | 0.80<br>(0.57-1.13) | .201 | 0.84<br>(0.58-1.20) | .340 |
| Medical mistrust | 0.98<br>(0.88-1.09) | .713 | 0.97<br>(0.86-1.09) | .581 | 0.93<br>(0.70-1.24) | .634 | 0.91<br>(0.67-1.24) | .548 |
| Trust in physician | 1.09<br>(0.96-1.25) | .171 | 1.07<br>(0.93-1.24) | .328 | 0.66<br>(0.43-1.03) | .069 | 0.71<br>(0.44-1.14) | .159 |
| Family loyalty | 1.12<br>(0.98-1.29) | .098 | 1.15<br>(0.98-1.35) | .088 | 1.28<br>(0.83-1.97) | .272 | 1.35<br>(0.82-2.20) | .239 |
| Any religious objection to LDKT<br>(Reference: No religious objection to LDKT) | 0.95<br>(0.82-1.12) | .563 | 0.93<br>(0.78-1.10) | .389 | 1.46<br>(0.92-2.31) | .110 | 1.37<br>(0.82-2.28) | .231 |
| Religiosity | 1.00<br>(0.97-1.03) | .826 | 0.98<br>(0.95-1.02) | .288 | 1.10<br>(1.01-1.21) | .036 | 1.10<br>(0.99-1.22) | .064 |
| Health literacy | 1.00<br>(0.93-1.08) | .979 | 1.00<br>(0.92-1.10) | .937 | 1.13<br>(0.84-1.52) | .415 | 1.09<br>(0.79-1.50) | .600 |
| *LASSO-selected covariates:* |  |  |  |  |  |  |  |  |
| Demographic characteristics |  |  |  |  |  |  |  |  |
| Age | - | - | - | - | **0.98**<br>**(0.96-1.00)** | **.020** | **0.98**<br>**(0.96-1.00)** | **.038** |
| Equivalent of 12<sup>th</sup> grade education or less<br>(Reference: More than 12<sup>th</sup> grade education) | - | - | - | - | **0.44**<br>**(0.25-0.76)** | **.003** | **0.41**<br>**(0.22-0.77)** | **.005** |
| Family Income of \$50,000 or less<br>(Reference: Greater than \$50,000) | 0.85<br>(0.71-1.01) | .064 | 0.87<br>(0.71-1.06) | .167 | **0.36**<br>**(0.22-0.58)** | **< .001** | **0.42**<br>**(0.25-0.70)** | **.001** |
|  | Intent-to-Treat |  | Per-Protocol |  | Intent-to-Treat |  | Per-Protocol |  |
|  | SHR<br>(95% CI) | p-value | SHR<br>(95% CI) | p-value | SHR<br>(95% CI) | p-value | SHR<br>(95% CI) | p-value |
| Separated, divorced, or widowed<br>(Reference: Married/partnered) | **0.84**<br>**(0.72-0.99)** | **.038** | **0.82**<br>**(0.69-0.98)** | **.028** | - | - | - | - |
| Employment (Unemployed)<br>(Reference: Part- or full-time employment) | 0.87<br>(0.74-1.04) | .119 | **0.80**<br>**(0.66-0.96)** | **.016** | 0.70<br>(0.43-1.12) | .137 | 0.67<br>(0.40-1.13) | .134 |
| Number in one's social network<br>*Note: log was taken to adjust for nonnormality* | **0.89**<br>**(0.81-0.97)** | **.011** | **0.85**<br>**(0.77-0.95)** | **.003** | - | - | - | - |
| Medical factors |  |  |  |  |  |  |  |  |
| Dialysis modality<br>(Reference: Not on dialysis) |  |  |  |  |  |  |  |  |
| Hemodialysis | 0.90<br>(0.53-1.53) | .708 | 0.78<br>(0.40-1.50) | .453 | - | - | - | - |
| Peritoneal dialysis | 0.86<br>(0.48-1.51) | .593 | 0.74<br>(0.37-1.47) | .388 | - | - | - | - |
| Dialysis duration<br>(Reference: No time on dialysis) |  |  |  |  |  |  |  |  |
| <1 year on dialysis | 0.82<br>(0.48-1.40) | .472 | 0.99<br>(0.51-1.92) | .972 | **0.48**<br>**(0.28-0.82)** | **.007** | **0.51**<br>**(0.29-0.90)** | **.020** |
| 1-5 years on dialysis | 0.87<br>(0.50-1.53) | .638 | 0.97<br>(0.49-1.93) | .941 | **0.39**<br>**(0.17-0.89)** | **.025** | **0.39**<br>**(0.16-0.97)** | **.043** |
| >5 years on dialysis | 0.62<br>(0.33-1.16) | .136 | 0.73<br>(0.34-1.56) | .414 | 0.38<br>(0.09-1.51) | .168 | 0.46<br>(0.10-2.06) | .310 |
| BMI<br>*Note: value was squared to adjust for nonnormality* | 1.00<br>(0.99-1.00) | .165 | 1.00<br>(0.34-1.56) | .414 | - | - | - | - |
| Psychosocial Characteristics |  |  |  |  |  |  |  |  |
| Moderate-to-severe Depression<br>(Reference: No depression) | **0.63**<br>**(0.45-0.89)** | **.009** | 0.73<br>(0.50-1.05) | .088 | - | - | - | - |
| Transplant Factors |  |  |  |  |  |  |  |  |
| Number of learning activities | 1.06<br>(0.99-1.13) | .070 | **1.08**<br>**(1.00-1.16)** | **.037** | - | - | - | - |
| Donor Recruitment and Preferences |  |  |  |  |  |  |  |  |
| Does not have a living donor at baseline<br>(Reference: Has a living donor at baseline) | - | - | - | - | **0.38**<br>**(0.24-0.62)** | **< .001** | **0.41**<br>**(0.25-0.67)** | **< .001** |

*Note:* <sup>a-d</sup> Bolded value indicate statistical significance of $p \leq .05$ . Cells marked with '-' indicate that the variable was not included in the model for that outcome (i.e., not selected by LASSO) and are intentionally left blank.
<sup>a</sup> **Evaluation Completion:** Intent-to-treat and Per-protocol samples, event counts (primary, censored, competing), and missing values are detailed in Supplemental Table 3.
<sup>b</sup> **LDKT Receipt:** Intent-to-treat and Per-protocol samples, event counts (primary, censored, competing), and missing values are detailed in Supplemental Table 4.

**Table 3.** Fine-Gray proportional sub-distribution hazards models, examining interaction effects ^a^ with TALK-Assignment and 9 pre-specified variables. Results display analyses in adjusted models on outcomes of evaluation completion (competing event: death) and LDKT receipt (competing events: DDKT, death). Intent-to-treat and Per-Protocol approaches are shown below.

| Variables | Evaluation Completion <sup>b</sup> |  |  |  | LDKT Receipt <sup>c</sup> |  |  |  |
| --- | --- | --- | --- | --- | --- | --- | --- | --- |
|  | Intent-to-Treat |  | Per-Protocol |  | Intent-to-Treat |  | Per-Protocol |  |
|  | SHR<br>(95% CI) | p-value<br>(corrected<br>p-value <sup>d</sup>) | SHR<br>(95% CI) | p-value<br>(corrected<br>p-value <sup>d</sup>) | SHR<br>(95% CI) | p-value<br>(corrected<br>p-value <sup>d</sup>) | SHR<br>(95% CI) | p-value<br>(corrected<br>p-value <sup>d</sup>) |
| Race and Ethnicity |  |  |  |  |  |  |  |  |
| TALK Assignment x<br>Race and ethnicity<br>(Reference: No-TALK,<br>Non-Hispanic White) |  |  |  |  |  |  |  |  |
| TALK x Non-Hispanic<br>Black | 0.72<br>(0.50-1.04) | .078 (.390) | 0.58<br>(0.38-0.88) | **.011** (.055) | 0.57<br>(0.14-2.29) | .429 (.858) | 0.69<br>(0.16-3.06) | .629 (.786) |
| TALK x Other race or<br>ethnicity | 0.73<br>(0.42-1.25) | .253 (.440) | 0.77<br>(0.43-1.40) | .393 (.655) | 0.91<br>(0.16-5.20) | .915 (.915) | 1.03<br>(0.10-10.09) | .983 (.992) |
| Experiences of Discrimination in Healthcare |  |  |  |  |  |  |  |  |
| TALK Assignment x<br>Experiences of<br>discrimination<br>(Reference: No-TALK,<br>No-discrimination) | 0.66<br>(0.48-0.90) | **.009** (.090) | 0.42<br>(0.29-0.61) | **< .001** (.001) | 1.75<br>(0.56-5.52) | .338 (.858) | 1.48<br>(0.43-5.07) | .530 (.786) |
| Perceived Racism in Healthcare |  |  |  |  |  |  |  |  |
| TALK Assignment x<br>Perceived Racism<br>(Reference: No-TALK) | 0.97<br>(0.82-1.15) | .721 (.721) | 0.85<br>(0.70-1.03) | .091 (.182) | 1.19<br>(0.64-2.20) | .579 (.915) | 1.17<br>(0.62-2.20) | .627 (.786) |
| Medical Mistrust |  |  |  |  |  |  |  |  |
| TALK Assignment x<br>Medical Mistrust<br>(Reference: No-TALK) | 0.90<br>(0.76-1.08) | .264 (.440) | 0.83<br>(0.68-1.00) | **.049** (.163) | 0.75<br>(0.45-1.24) | .263 (.858) | 0.69<br>(0.39-1.20) | .183 (.786) |
| Trust in Physician |  |  |  |  |  |  |  |  |
| TALK Assignment x<br>Trust in Physician<br>(Reference: No-TALK) | 1.09<br>(0.85-1.40) | .476 (.680) | 1.11<br>(0.84-1.45) | .461 (.659) | 0.92<br>(0.41-2.05) | .834 (.915) | 0.80<br>(0.33-1.96) | .626 (.786) |
| Family Loyalty |  |  |  |  |  |  |  |  |
| TALK Assignment x<br>Family Loyalty<br>(Reference: No-TALK) | 0.84<br>(0.65-1.09) | .187 (.440) | 0.76<br>(0.57-1.02) | .066 (.165) | 1.62<br>(0.71-3.72) | .254 (.858) | 1.52<br>(0.56-4.14) | .414 (.786) |
| Religious Objection to LDKT |  |  |  |  |  |  |  |  |
| TALK Assignment x<br>Religious Objection to<br>LDKT<br>(Reference, No-TALK,<br>No religious objection) | 1.07<br>(0.79-1.44) | .679 (.721) | 0.99<br>(0.71-1.37) | .930 (.930) | 1.23<br>(0.50-3.00) | .653 (.915) | 1.36<br>(0.51-3.66) | .541 (.786) |
| Religiosity |  |  |  |  |  |  |  |  |
| TALK Assignment x<br>Religiosity<br>(Reference: No-TALK) | 1.04<br>(0.99-1.11) | .135 (.440) | 1.01<br>(0.94-1.07) | .833 (.930) | 1.02<br>(0.85-1.22) | .824 (.915) | 1.00<br>(0.82-1.22) | .992 (.992) |
|  | Intent-to-Treat |  | Per-Protocol |  | Intent-to-Treat |  | Per-Protocol |  |
|  | SHR<br>(95% CI) | *p*-value<br>(corrected<br>*p*-value <sup>d</sup>) | SHR<br>(95% CI) | *p*-value<br>(corrected<br>*p*-value <sup>d</sup>) | SHR<br>(95% CI) | *p*-value<br>(corrected<br>*p*-value <sup>d</sup>) | SHR<br>(95% CI) | *p*-value<br>(corrected<br>*p*-value <sup>d</sup>) |
| Health Literacy |  |  |  |  |  |  |  |  |
| TALK Assignment x<br>Health Literacy<br>(Reference: No-TALK) | 0.97<br>(0.85-1.11) | .675 (.721) | 1.01<br>(0.86-1.18) | .904 (.930) | 0.77<br>(0.44-1.37) | .382 (.858) | 0.84<br>(0.45-1.56) | .580 (.786) |

Note: <sup>a-d</sup> Bolded *p*-values denotes statistical significance of $p < .05$ . Interaction terms are shown relative to reference categories (e.g., No-TALK and No-Discrimination).
<sup>a</sup> Each interaction term (i.e., TALK-assignment x key variable) represents nine separate adjusted models. Each model included the same set of covariates: the key variable x TALK, the remaining 8 pre-specified variables not included in the interaction term, and the LASSO-selected variables (see Table 2 for main effects and list of all variables).
<sup>b</sup> **Evaluation Completion:** Intent-to-treat and Per-protocol samples for adjusted model, event counts (primary, censored, competing), and missing values are detailed in Supplemental Table 3.
<sup>c</sup> **LDKT Receipt:** Intent-to-treat and Per-protocol samples for adjusted model, event counts (primary, censored, competing), and missing values are detailed in Supplemental Table 4.
<sup>d</sup> Correction for false discovery rate was conducted using the Benjamini-Hochberg method, that adjusts these values by accounting for both number of comparisons and rank (Adjusted *p*-value = original *p*-value \* total number of tests / rank). Adjusted *p*-values are interpreted according to conventional significance thresholds ( $p < .05$ ).

#### TALK Intervention Effects in Adjusted Modeling

*Evaluation completion:* We identified no difference in the cumulative incidence of evaluation completion by TALK-assignment (TALK versus No-TALK SHR =1.06; 95% CI: 0.92-1.22; *p*=.408). Findings were consistent across intent-to-treat and per-protocol approaches (Table 2, Figure 2A).

**Figure 2.**
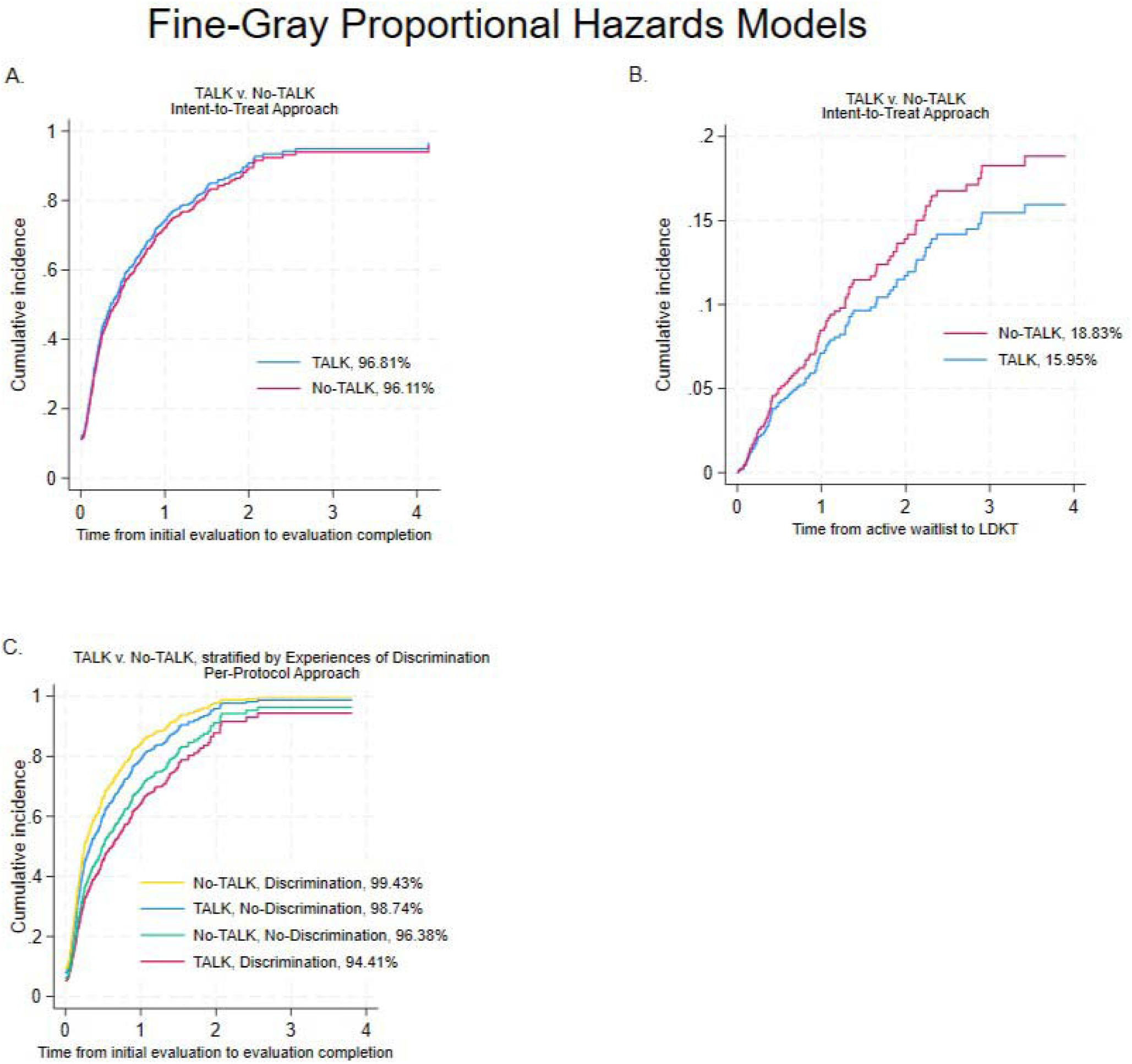
Fine-Gray proportional hazard models using cumulative incidence function: A. KT Evaluation Completion by TALK-Assignment (Main Effects, Intent-to-Treat); B. LDKT Receipt by TALK-Assignment (Main Effects, Intent-to-Treat); C. KT evaluation completion by TALK-assignment and Experiences of Discrimination (Per-Protocol)

*LDKT receipt:* We identified no difference in the cumulative incidence of LDKT receipt by TALK-assignment (SHR=0.83; 95% CI: 0.55-1.24; *p*=.380). Findings were consistent across intent-to-treat and per-protocol approaches (Table 2, Figure 2B).

### Exploring Interaction Effects of the TALK Intervention

*Evaluation completion:* Across the nine pre-specified key variables, only one interaction with TALK-assignment remained statistically significant after correcting for multiple comparisons (Table 3). Under the per-protocol approach, we identified a significant interaction between TALK-assignment and experiences of healthcare-related discrimination (i.e., “Discrimination” versus “No-Discrimination”) on the cumulative incidence of completing evaluation (SHR=0.42; 95% CI: 0.29-0.61; corrected *p*=.001). This effect was not significant under the intent-to-treat approach following correction for multiple comparisons, despite trending in the same direction (SHR=0.66; 95% CI: 0.48-0.90; corrected *p*=.090).

To interpret the interaction, we examined simple effects of TALK-assignment at each level of discrimination, computed as linear contrasts of the regression coefficients. Within the “No-Discrimination” group, TALK participants had a significantly higher cumulative incidence of evaluation completion than No-TALK participants (SHR=1.32, 95% CI: 1.10-1.58, *p*=.003; 98.7% versus 96.4% evaluation completion rate). Within the “Discrimination” group, TALK participants had a significantly lower cumulative incidence of evaluation completion than No-TALK participants (SHR=0.56, 95% CI: 0.41-0.77, *p*<.001; 94.4% versus 99.4% evaluation completion rate). See Figure 2C and Table 4.

**Table 4.** Contrast effects demonstrating differences by TALK-assignment (TALK v. No-TALK) and Experiences of Discrimination (Discrimination v. No-Discrimination) for the cumulative incidence of evaluation completion under Per-Protocol approach.

|  | No-Discrimination |  | Discrimination |  |
| --- | --- | --- | --- | --- |
|  | SHR<br>(95% CI) | *p*-value | SHR<br>(95% CI) | *p*-value |
| TALK (Reference: No-TALK) | 1.32<br>(1.10-1.58) | **.003** | 0.56<br>(0.41-0.77) | **< .001** |

*Note:* Bolded *p*-values denotes statistical significance of $p < .05$ . Figure 2C displays these relationships.

*LDKT receipt:* We found no significant interactions between TALK-assignment and the nine pre-specified key variables on the cumulative incidence of LDKT receipt. Findings were consistent across intent-to-treat and per-protocol approaches (Table 3).

## Discussion

Our RCT, conducted under conditions of gold-standard care,^19,20^ determined that the education-only component of the TALK intervention did not increase evaluation completion or LDKT receipt. Although we hypothesized that introducing TALK materials at KT evaluation initiation would improve outcomes without resource-intensive components, our results suggest education materials did not drive significant change in this context.

In exploratory analyses, we identified one significant, yet counterintuitive, interaction between TALK-assignment and experiences of healthcare discrimination, when predicting time to evaluation completion, observed only under the per-protocol approach. Although participants were randomized, this per-protocol sample reflected self-reported engagement with TALK materials rather than assignment alone, while the No-TALK group remained unchanged from the intent-to-treat analysis. The interaction remained statistically significant even after correction for multiple comparisons. Although TALK was associated with greater evaluation completion among participants who had not experienced discrimination in healthcare, the opposite pattern was observed among those who *had* experienced prior healthcare-related discrimination, thus underscoring the relevance of measuring discrimination among patients receiving an educational intervention such as TALK. This exploratory analysis sheds light on differential effects within the TALK arm, and suggests that patients’ past healthcare-related discrimination experiences may limit the impact of educational interventions.

The present findings did not support our hypothesis that TALK materials alone would be sufficient to promote patient engagement that subsequently improved KT outcomes, even if presented earlier in the KT process, and within a context where barriers to completing evaluation were minimized. This is consistent with prior studies,^22,24^ suggesting that TALK is unlikely to meaningfully influence complex health outcomes such as evaluation completion or LDKT receipt. Although high rates of evaluation completion within the context of KTFT may have limited the potential for additional improvements in that outcome, the absence of an effect on LDKT receipt (i.e., an outcome shaped by clinical, social, and systemic factors^7,12,13,18^) suggests that interventions focused solely on patient education are unlikely to influence downstream KT-related outcomes. These findings highlight a potential misalignment between multi-level factors relevant to LDKT, and the individual-level focus of stand-alone educational strategies.

Presenting TALK materials without any structured guidance on how to review them may have inadvertently burdened patients, particularly those with a history of prior healthcare-related discrimination. Essentially, patients were expected to translate information into action while already navigating complex LDKT-related decision-making during the evaluation process.

Greater clinic support such as requiring physicians/staff to discuss the materials with patients before their initial evaluation appointment in a standardized fashion might have removed the burden from patients to review materials independently. Such an approach may have increased saliency of topics covered, while also providing patients with more direction on how to proceed with the new information. Unfortunately, this approach requires greater personnel involvement, highlighting the challenges of implementing a scaled-back version of TALK that produces observable changes in lower-resourced settings. Overall, these null findings underscore the limits of stand-alone educational strategies even within high-quality care conditions.

Our findings point to the potential value of designing interventions that shift responsibilities away from patients, and toward broader, system-level support—ideally in ways that are scalable, and do not overload clinic resources. Moving forward, prioritizing comprehensive healthcare system changes that equitably serve all patients may be more effective than relying on patients to act on knowledge gained from education materials. For example, initiatives like KTFT^20^ or utilizing health system surveillance registries to provide early detection of barriers to LDKT among KT candidates,^50,51^ may be more sustainable and effective alternatives.

### Limitations and Strengths

This study was conducted within one KT center, thus limiting generalizability of findings. The TALK intervention was delivered within the KTFT cohort,^19^ a streamlined evaluation approach associated with increased KT waitlisting,^20^ leaving little room for TALK to make additional improvements toward evaluation completion. Consequently, the study context may have limited the observable impact of a scaled-back educational intervention.

Our power calculation was based on LDKT receipt as a binary outcome. Although evaluation completion, and exploratory interaction and per-protocol analyses were not included in pre-trial power calculations, the sample size and use of Fine-Gray competing risk survival modeling allowed for inclusion of censored cases, thus providing sufficient data to meaningfully assess these outcomes.

Ultimately, this study included a diverse sample from a large center that serves urban and rural populations. We were the first to test TALK starting at evaluation initiation, we tested a scalable intervention to assess potential for broader implementation, and we conducted both intent-to-treat and per-protocol analyses, yielding a rigorous evaluation of the TALK intervention’s impact.

### Conclusions

Our RCT yielded null findings under gold-standard care conditions. In this trial, TALK education materials presented to patients initiating KT evaluation did not increase evaluation completion or LDKT receipt. Healthcare-related discrimination influenced intervention outcomes only within the per-protocol analysis, underscoring the importance of considering sociocultural factors when implementing educational interventions. These results come from a large, well-powered trial with long-term follow-up, offering robust evidence that a stand-alone educational approach is unlikely to meaningfully improve KT-related outcomes. Although TALK did not improve KT-related outcomes, another study demonstrated that streamlining the evaluation process improved waitlisting and KT receipt within this same sample.^20^ Future studies may consider prioritizing healthcare system interventions, instead of educational interventions, to promote LDKT among patients completing KT evaluations.

## Acknowledgment Section

### Disclosure of potential conflicts of interest

None to disclose.

### Funding/Support

This work was supported in part by grants R01DK081325 from the National Institute of Diabetes and Digestive and Kidney Diseases (PI: L. Myaskovsky); UL1TR001857 from the National Center for Advancing Translational Sciences in part to Dr. Myaskovsky (PI: S. Reis); T32HL007736 from the National Heart, Lung, and Blood Institute in part to Dr. Vélez-Bermúdez (PI: T. Resta); and C-3924 from Dialysis Clinic Inc (PI: L. Myaskovsky).

### Role of the Funder/Sponsor

The funders had no role in the design and conduct of the study; collection, management, analysis, and interpretation of the data; preparation, review, or approval of the manuscript; and decision to submit the manuscript for publication.

### Data Sharing Statement

Dr. Larissa Myaskovsky had full access to all the data in the study and takes responsibility for the integrity of the data and the accuracy of the data analysis. The data are not publicly available due to privacy or ethical restrictions.

## Author Contributions

Dr. Vélez-Bermúdez is the first author. Dr. Myaskovsky is the senior author.

*Concept and design:* Vélez-Bermúdez, Loor, Dew, Boulware, and Myaskovsky.

*Acquisition, analysis, or interpretation of data*: Vélez-Bermúdez, Loor, Leyva, Zhu, Croswell, Unruh, Dew, and Myaskovsky.

*Drafting of the manuscript:* Vélez-Bermúdez and Myaskovsky.

*Critical review of the manuscript for important intellectual content:* All authors.

*Statistical analysis*: Vélez-Bermúdez, Leyva, Zhu.

*Obtained funding*: Myaskovsky.

*Supervision:* Myaskovsky, Zhu, Unruh, and Dew.

## Acknowledgments

The authors thank the individuals who participated in this study for their contributions to this research.

## Notes

### Competing Interest Statement

The authors have declared no competing interest.

### Clinical Trial

NCT02342119

### Clinical Protocols

https://doi.org/10.1016/j.cct.2016.11.011

### Author Declarations

The Institutional Review Boards at the University of Pittsburgh (PRO09060113) and the University of New Mexico (17-084) gave ethical approval for this work.

