## Supplemental Material for "Education Intervention for Evaluation and Living Donor Kidney Transplantation: A randomized trial"

**Supplement Contents**

**Supplemental Table 1.** Consolidated Standards of Reporting Trials Checklist

**Supplemental Table 2.** Baseline characteristics and potential predictors

**Supplemental Table 3.** Evaluation Completion: Primary, Censored, and Competing Event Counts by Analytic Approach and Model Adjustment

**Supplemental Table 4.** LDKT Receipt: Primary, Censored, and Competing Event Counts by Analytic Approach and Model Adjustment

**Supplemental References.**

**Supplemental Table 1.** Consolidated Standards of Reporting Trials Checklist ^1^

| Section/Topic | Item No | Checklist item | Reported on page No |
| --- | --- | --- | --- |
| Title and abstract | | | |
|  | 1a | Identification as a randomized trial in the title | 1 |
|  | 1b | Structured summary of trial design, methods, results, and conclusions (for specific guidance see CONSORT for abstracts) | 2-4 |
| Introduction | | | |
| Background and objectives | 2a | Scientific background and explanation of rationale | 5-7 |
|  | 2b | Specific objectives or hypotheses | 6-7 |
| Methods | | | |
| Trial design | 3a | Description of trial design (such as parallel, factorial) including allocation ratio | 7-8 |
|  | 3b | Important changes to methods after trial commencement (such as eligibility criteria), with reasons | 8-9 |
| Participants | 4a | Eligibility criteria for participants | 7 |
|  | 4b | Settings and locations where the data were collected | 7 |
| Interventions | 5 | The interventions for each group with sufficient details to allow replication, including how and when they were actually administered | 7-8 |
| Outcomes | 6a | Completely defined pre-specified primary and secondary outcome measures, including how and when they were assessed | 8 |
|  | 6b | Any changes to trial outcomes after the trial commenced, with reasons | N/A |
| Sample size | 7a | How sample size was determined | 8 |
|  | 7b | When applicable, explanation of any interim analyses and stopping guidelines | N/A |
| Randomization: |  |  |  |
| Sequence generation | 8a | Method used to generate the random allocation sequence | 8 |
|  | 8b | Type of randomization; details of any restriction (such as blocking and block size) | 8 |
| Allocation concealment mechanism | 9 | Mechanism used to implement the random allocation sequence (such as sequentially numbered containers), describing any steps taken to conceal the sequence until interventions were assigned | 8 |
| Implementation | 10 | Who generated the random allocation sequence, who enrolled participants, and who assigned participants to interventions | 8 |
| Blinding | 11a | If done, who was blinded after assignment to interventions (for example, participants, care providers, those assessing outcomes) and how | 8 |
|  | 11b | If relevant, description of the similarity of interventions | N/A |
| Statistical methods | 12a | Statistical methods used to compare groups for primary and secondary outcomes | 9-12 |
|  | 12b | Methods for additional analyses, such as subgroup analyses and adjusted analyses | 10-12 |
| Results | | | |
| Participant flow (a diagram is strongly recommended) | 13a | For each group, the numbers of participants who were randomly assigned, received intended treatment, and were analyzed for the primary outcome | 12, Table 1, Figure 1, Supplemental Tables 3-4 |
|  | 13b | For each group, losses and exclusions after randomization, together with reasons | Figure 1, Supplemental Tables 3-4 |
| Recruitment | 14a | Dates defining the periods of recruitment and follow-up | 8 |
|  | 14b | Why the trial ended or was stopped | 7 |
| Baseline data | 15 | A table showing baseline demographic and clinical characteristics for each group | Table 1 |
| Numbers analyzed | 16 | For each group, number of participants (denominator) included in each analysis and whether the analysis was by original assigned groups | 12, Figure 1, Supplemental Tables 3-4 |
| Outcomes and estimation | 17a | For each primary and secondary outcome, results for each group, and the estimated effect size and its precision (such as 95% confidence interval) | Tables 2-3 |
|  | 17b | For binary outcomes, presentation of both absolute and relative effect sizes is recommended | N/A |
| Ancillary analyses | 18 | Results of any other analyses performed, including subgroup analyses and adjusted analyses, distinguishing pre-specified from exploratory | Table 2-3 |
| Harms | 19 | All important harms or unintended effects in each group (for specific guidance see CONSORT for harms) | N/A |
| Discussion | | | |
| Limitations | 20 | Trial limitations, addressing sources of potential bias, imprecision, and, if relevant, multiplicity of analyses | 14-17 |
| Generalizability | 21 | Generalizability (external validity, applicability) of the trial findings | 14-17 |
| Interpretation | 22 | Interpretation consistent with results, balancing benefits and harms, and considering other relevant evidence | 14-17 |
| Other information | | |  |
| Registration | 23 | Registration number and name of trial registry | 4, 7 |
| Protocol | 24 | Where the full trial protocol can be accessed, if available | 4, 7 |
| Funding | 25 | Sources of funding and other support (such as supply of drugs), role of funders | 18 |

| **Supplemental Table 2.** **Baseline characteristics, independent variables, and potential covariates** | | | |
| --- | --- | --- | --- |
| **Variables** | | **Description** | **Coding/ Range/ Cronbach’s α (if applicable)** |
| **Demographic, sociocultural, and psychosocial factors at baseline included in all multivariable analyses** | | | |
|  | Race and ethnicity | Grouped into the following categories:  1. Non-Hispanic White  2. Non-Hispanic Black  3. Other (includes American Indian or Alaska Native, Asian, Hispanic or Latine, Native Hawaiian or Pacific Islander, or Mixed race or Other) | Categorical, 3 categories |
|  | Experience of discrimination in healthcare settings^2,3^ | 7 items assessing perceived discrimination in a healthcare setting; original range: 1 (never) to 5 (always) | Dichotomized for analysis: “ever experienced discrimination” versus “never experienced discrimination”  Cronbach’s α = 0.88 |
|  | Perceived racism in healthcare settings^4,5^ | 4 items assessing patients’ belief that racism is common in healthcare | Mean score calculated for analysis  Continuous  Responses and potential range: 1 (strongly disagree) to 5 (strongly agree)  Cronbach’s α = 0.75 |
|  | Medical mistrust^5,6^ | 7 items assessing beliefs that patient’s hospital is trustworthy, competent, and acting in their best interests | Mean score calculated for analysis  Continuous  Responses and potential range: 1 (strongly disagree) to 5 (strongly agree);  Cronbach’s α = 0.78 |
|  | Trust in physicians^7^ | 11 items assessing patients’ trust in their physician | Mean score calculated for analysis  Continuous  Responses and potential range: 5 (totally disagree) to 1 (totally agree)  Cronbach’s α = 0.85 |
|  | Family loyalty^8^ | 16 items assessing participants’ loyalty and mutual support regarding the family | Mean score calculated for analysis  Continuous  Responses and potential range: 1 (totally disagree) to 5 (totally agree);  Cronbach’s α = 0.84 |
|  | Overall Religiosity^9^ | 2 items assessing the importance and influence of religious beliefs in a participants’ life | Mean score calculated for analysis  Continuous  Responses and potential range: 1 (not at all important) to 9 (very important);  Cronbach’s α = 0.82 |
|  | Religious objections to LDKT^10^ | Revised subscale of the 8 item Organ Donation Attitude Survey (ODAS). From these questions, we categorized respondents into 3 groups:  1. No objection – “disagree” or “strongly disagree” with all religious objections to transplant  2. Neutral – combination of “disagree,” “strongly disagree” and “not sure” toward religious objection to transplant  3. Any objection – “agree” or “strongly agree” with any religious objection to transplant | Dichotomized for analysis: “any religious objection to LDKT” versus “no religious objection to LDKT”  Cronbach’s α = 0.72 |
|  | Health Literacy^11^ | A three-item measure assessing participants’ understanding about their own health information, with scores ranging from 1 (“Always,” “Extremely”) to 5 (“Never,” “Not at all”). Higher scores reflect higher literacy. The measure was scored by taken the mean score of each item. In this sample, health literacy scores ranged from 1 – 5. | Mean score calculated for analysis  Continuous  Cronbach’s α = 0.67 |
| **Demographic characteristics, tested in LASSO for inclusion in subsequent multivariable analyses** | | | |
|  | Age | Age at baseline interview | Continuous  Range in present sample: 20 – 88 |
|  | Gender | Men, women | Dichotomous |
|  | Educational level | Patient-reported using the following choices:  1. Less than high school  2. Some high school  3. High school graduate  4. Some college  5. College degree  6. Graduate degree | Dichotomized for analysis: “Less than or equal to high school” versus “Greater than high school education” |
|  | Family Income | Patient-reported using the following choices:  1. Under $15,000 2. $15,000 - $24,999 3. $25,000 - $49,999 4. $50,000 - $74,999 5. $75,000 - $100,000 6. Over $100,000 | Dichotomized for analysis: “Less than $50,000” versus “Greater than or equal to $50,000” |
|  | Marital status | Patient-reported using the following categories:  1. Single (never married)  2. Separated or Divorced  3. Widowed  4. Married  5. Domestic Partnership | Dichotomized for analysis: “Married/In a Domestic partnership” versus “Single/Separated or Divorced/Widowed” |
|  | Insurance status | Patient-reported and grouped into the following categories:  1. Private insurance  2. Private/public mix  3. Public insurance only | Categorical, 3 categories |
|  | Employment status | Patient-reported using the following categories:  1. Yes, full time  2. Yes, part time  3. No, unemployed | Dichotomized for analysis: “Full- or part-time employment” versus “unemployed” |
|  | Number in social network (i.e., “network of potential living donors”) | Patient-reported number of the network of potential living donors available for evaluation was determined by asking participants to indicate how many living relatives and friends they had aged 18–70 years of age. The log value was calculated to adjust for skewness. | Continuous  Range in present sample: 0 – 150 |
| **Medical factors, tested in LASSO for inclusion in subsequent multivariable analyses** | | | |
|  | Body mass index (BMI) | Calculated from medical record, value was squared in analysis to adjust for skewness | Continuous  Range in present sample: 13.49 – 53.84 |
|  | Charlson Comorbidity Index^12^ | From medical record data: weighted score reflecting the number and severity of co-morbid health conditions | Continuous: 0 (no comorbidities) to 33 (a higher number of comorbidities or more serious comorbidities)  Range in present sample: 2 – 11 |
|  | Dialysis duration | From medical record:  1. 0 years on dialysis  2. <1 year on dialysis  3. 1-<5 years on dialysis  4. > 5 years | Categorical, 4 categories |
|  | Dialysis type | From medical record: 1. Hemodialysis  2. Peritoneal dialysis  3. No dialysis | Categorical, 3 categories |
|  | Burden of Kidney Disease^13^ | Subscale that measures the extent to which kidney disease interferes with the respondent’s life. | Mean score calculated for analysis  Continuous  Responses and potential range: 1 (Definitely false) to 5 (Definitely true)  Cronbach’s α = 0.76 |

| **Psychosocial characteristics, tested in LASSO for inclusion in subsequent multivariable analyses** | | | | | |
| --- | --- | --- | --- | --- | --- |
|  | Social support^14,15^ | | 12-item Interpersonal Support Evaluation List (ISEL-12) - assessed perceived availability of 3 separate functions of social support: "tangible," "appraisal,” and "belonging". | | Continuous  Range in present sample: 12 – 48  Cronbach’s α = 0.86 |
|  | Anxiety ^16^ | | 6-item Brief Symptom Inventory (BSI) Anxiety subscale where participants report the extent to which they indicate how bothered or distressed they have felt in the past 2 weeks by several symptoms. Responses ranged from 1 (“Not at all”) to 5 (“Extremely”). Responses were dichotomized to reflect “No anxiety” or “Moderate to severe anxiety.” | | Dichotomized for analysis: “No anxiety” versus “Moderate to severe anxiety”  Cronbach’s α = 0.83 |
|  | Depression ^16^ | | 6-item Brief Symptom Inventory (BSI) Depression subscale where participants report the extent to which they indicate how bothered or distressed they have felt in the past 2 weeks by several symptoms. Responses ranged from 1 (“Not at all”) to 5 (“Extremely”). Responses were dichotomized to reflect “No depression” or “Moderate to severe depression.” | | Dichotomized for analysis: “No depression” versus “Moderate to severe depression”  Cronbach’s α = 0.83 |
| **Transplant knowledge and concerns, tested in LASSO for inclusion in subsequent multivariable analyses** | | | | | |
|  | Transplant knowledge^17^ | Participants were assessed on their knowledge about transplant using a 19-item KT Knowledge Survey. Items 1 – 8 were multiple choice, and items 9 – 19 were “True” or “False.” Each item answered correctly adds 1 to total score. | | Continuous  Range in present sample: 0 – 18  Cronbach’s α = 0.84 | |
|  | Number of learning activities^18^ | Participants reported the type and number of KT-related learning activities (e.g., reading brochures, online research) with greater numbers indicating engagement in more learning activities. | | Continuous  Range in present sample: 0 – 4  Cronbach’s α = 0.54 | |
|  | Hours engaged in learning activities^18^ | Participants reported the amount of time spent in KT-related learning activities (e.g., reading brochures, online research). | | Categorized for present analysis:  1: 0 – 2 hours of learning  2: Greater than 2 hours, less than or equal to 5 hours  3: Greater than 5 hours of learning  Cronbach’s α = 0.59 | |
|  | Transplant concerns | In a 24-item assessment, participants reported which of 24 common transplant-related concerns were most important in influencing their decision to pursue transplant. | | Sum score; continuous.  Item responses ranged from 1 (“Not important”) to 5 (“Extremely important”) Range in present sample: 16 – 60  Cronbach’s α = 0.80 | |
| **Donor recruitment and preference variables, tested in LASSO for inclusion in subsequent multivariable analyses** | | | | | |
|  | Donation preference | Patient-reported using the following categories:  1 = Transplant from a deceased donor  2 = Transplant from a living donor | | Dichotomous | |
|  | Having a living donor at baseline | Patient-reported using the following categories:  0 = No  1 = Yes | | Dichotomous | |
|  | Willingness to accept a living donor | Patient-reported using the following categories:  0 = No  1 = Yes | | Dichotomous | |
|  | Willingness to ask for a living donor | Patient-reported using the following categories:  0 = No  1 = Yes | | Dichotomous | |

**Supplemental Table 3. Evaluation Completion: Primary, Censored, and Competing Event Counts by Analytic Approach and Model Adjustment**

| **Evaluation Completion** | | | | | |
| --- | --- | --- | --- | --- | --- |
| Analytic Approach & Model | N | Primary event  (evaluation completion) | Censored | Competing events | Excluded due to missing values |
| Intent-to-Treat, Unadjusted | Total=1108 TALK=574  No-TALK=534 | 810 (accepted=495,  rejected = 315) | 275 (incomplete=253; patient withdrew=18; evaluation ongoing=4) | 23 (deceased) | - |
| Intent-to-Treat,  Adjusted | Total=1048 TALK=544  No-TALK=504 | 761  (accepted=472, rejected=289) | 266 (incomplete=245; withdrew=17; ongoing=4) | 21 (deceased) | 60 |
| Per-Protocol,  Unadjusted | Total=892  TALK=358  No-TALK=534 | 662 (accepted=447, rejected=215) | 221 (incomplete=208; withdrew=10; ongoing=3) | 9 (deceased) | - |
| Per-Protocol,  Adjusted | Total=847  TALK=343  No-TALK=504 | 625 (accepted=427, rejected=198) | 214 (incomplete=201; withdrew=10; ongoing=3) | 8 (deceased) | 45 |

*Note*: Per-Protocol sample includes participants assigned to TALK who self-reported reviewing either the booklet or video.

**Supplemental Table 4. LDKT Receipt: Primary, Censored, and Competing Event Counts by Analytic Approach and Model Adjustment**

| **LDKT Receipt** | | | | | |
| --- | --- | --- | --- | --- | --- |
| Analytic Approach & Model | N | Primary event  (LDKT Receipt) | Censored | Competing events | Excluded due to missing values |
| Intent-to-Treat, Unadjusted | Total=473 TALK=239  No-TALK=234 | 108 | 142 (still on waitlist=46; removed from waitlist=96) | 223 (DDKT receipt=169; deceased=54) | - |
| Intent-to-Treat,  Adjusted | Total=450 TALK=227  No-TALK=223 | 99 | 137 (still on waitlist=45; removed from waitlist=92) | 214 (DDKT receipt=160; deceased=54) | 23 |
| Per-Protocol,  Unadjusted | Total=426  TALK=192  No-TALK=234 | 94 | 130 (still on waitlist=43; removed from waitlist=87) | 202 (DDKT receipt=156; deceased=46) | - |
| Per-Protocol,  Adjusted | Total=406  TALK=183  No-TALK=223 | 86 | 126 (still on waitlist=42; removed from waitlist=84) | 194 (DDKT receipt=148; deceased=46) | 20 |

*Note*: Per-Protocol sample includes participants assigned to TALK who self-reported reviewing either the booklet or video.
